# Environmental air monitoring in international airports: A novel approach for enhanced pathogen surveillance

**DOI:** 10.1101/2025.09.22.25336185

**Authors:** Dawn Gratalo, Cindy R. Friedman, Valerie J. Morley, Xueting Qiu, Ian Ruskey, Andrew P. Rothstein, Patrick B. Tiburcio, Casandra W. Philipson, Thomas W.S. Aichele, Stephen M. Bart, Dustin Jaynes, Birgitte B. Simen, Shelby L O’Connor, David H. O’Connor

**Affiliations:** Ginkgo Biosecurity, 27 Drydock Ave 8th Floor, Boston, MA, USA; Division of Global Migration Health, Centers for Disease Control and Prevention, Atlanta, Georgia, USA; Department of Pathology and Laboratory Medicine, University of Wisconsin-Madison, Madison, WI USA; Former Affiliation; Department of Public Safety, Dallas/Fort Worth International Airport, Texas, USA

**Keywords:** Air monitoring, pathogen surveillance, airports, wastewater, SARS-CoV-2, influenza, metagenomic sequencing, environmental surveillance, pandemic preparedness, biosecurity

## Abstract

Early detection of outbreaks and emerging pathogens is critical for public health and global biosecurity. Airports, as major international travel hubs with dense, enclosed populations, are high-risk settings for disease transmission and potential pathogen introduction. The U.S. Centers for Disease Control and Prevention, in collaboration with Ginkgo Biosecurity and the University of Wisconsin–Madison, implemented air monitoring for pathogen surveillance in congregate areas at four U.S. international airports. From October 2023 to August 2024, SARS-CoV-2 was detected by PCR in 98.3% of air samples and influenza A in 17.2%. These results correlated with positivity trends from other sample modalities, including aviation wastewater, traveler nasal swabs, and national clinical surveillance data. Targeted amplicon sequencing of SARS-CoV-2 from air samples correlated with contemporaneous lineages in wastewater collected and sequenced from the same airports. Metagenomic enrichment sequencing detected 30 viral species and recovered high-quality genomes for SARS-CoV-2, influenza, bocavirus, and seasonal coronaviruses. Together, these findings demonstrate that air sampling is a complementary surveillance modality to aviation wastewater for early pathogen detection at ports of entry.

## Introduction

Epidemic and pandemic preparedness relies heavily on robust and adaptive surveillance systems capable of early pathogen detection. Global travel hubs, particularly international airports, are critical interfaces for the introduction and spread of emerging and re-emerging pathogens due to their dense, transient populations and high interconnectedness.^1–4^ Detecting pathogens and identifying new variants in airports is especially important, as illustrated during the COVID-19 pandemic, which underscored the urgent need for near real-time genomic surveillance strategies.^5^ While testing of nasal swabs from arriving travelers provides one modality for monitoring, the process can be labor-intensive and relies on traveler willingness to participate.

Environmental surveillance has emerged as a non-invasive approach to characterize pathogens circulating in communities. Wastewater surveillance, including community and aviation wastewater, has demonstrated utility in tracking both the prevalence and genetic diversity of viruses such as SARS-CoV-2.^6–8^ Air monitoring serves as a complementary public health surveillance tool, enabling the collection of pathogenic genetic material shed by individuals. This approach is particularly effective for capturing respiratory pathogens from diverse populations, including asymptomatic or pre-symptomatic individuals who might not otherwise engage with clinical testing or contribute to accessible wastewater systems. Studies have shown the effectiveness of air sampling in multiple congregate settings. In hospitals, airborne pathogen detection has correlated strongly with clinical data ^9^, and airborne pathogen genetic material has also been detected in other communal spaces such as schools.^10–12^ As a scalable and cost-effective surveillance option, non-invasive air monitoring is particularly relevant for high-traffic environments, such as international airports, that serve as gateways into communities.

While PCR-based detection confirms the presence of pathogen genetic material, genomic sequencing is essential for identifying specific lineages, tracking variant evolution, and informing the development of diagnostics and therapeutics.^13–16^ However, obtaining high-quality genomic data from air samples presents challenges due to low biomass and nucleic acid degradation, especially for RNA viruses.^17^ Advanced sequencing methodologies are therefore crucial. Targeted amplicon sequencing offers high sensitivity but is limited to a narrow range of pathogens, while metagenomic approaches provide an agnostic view but often lack sensitivity for low-abundance targets. Metagenomic enrichment sequencing represents a promising strategy to improve both breadth and sensitivity by enriching viral nucleic acids prior to sequencing.^9,18^

In this study, air monitoring surveillance was implemented in congregate areas of four major U.S. international airports to assess the feasibility of detection of SARS-CoV-2 and Influenza A virus genetic material in an international airport setting; evaluate the effectiveness of sequencing methods for viral genome characterization from these air samples; and determine the complementary value of air monitoring within existing pathogen surveillance frameworks. This study explores the development of air monitoring as a proactive biosecurity system, to enhance our capacity for early threat identification at the critical intersection of global travel and community health.

## Methods

ThermoFisher AerosolSense™ active air samplers were placed in congregate areas adjacent to customs and immigration inspection areas and within proximity of several hundred arriving international passengers at San Francisco International Airport, California (SFO); Newark Liberty International Airport, New Jersey (EWR); Washington Dulles International Airport, Virginia (IAD); and Dallas/Fort Worth International Airport, Texas (DFW). Samplers were placed on tables 28-30 inches above the ground. The ThermoFisher AerosolSense™ has a built-in pump, that operates at 200 liters per minute, and a proprietary sample collection cartridge. Each ThermoFisher AerosolSense™ unit collected samples for 10 hours per day. ThermoFisher AerosolSense™ cartridges containing polyurethane foam substrates were removed from each air sampling machine after approximately 20 hours of use Monday through Thursday and after 30 hours of use Friday through Sunday.

Samples were collected from October 18, 2023–August 27, 2024, at SFO, EWR, and IAD, and from February 7–August 14, 2024, at DFW. At SFO, a second sampler was added on June 3, 2024 (denoted by the orange star in Figure 1), and both instruments operated until study end. Cartridges were packaged and shipped at room temperature to UW-Madison for extraction and qRT-PCR analyses. All results were linked to airport and collection date via unique barcodes. Each substrate was submerged in 500 µL of 1X phosphate-buffered saline (PBS) and vortexed.

**Figure 1:**
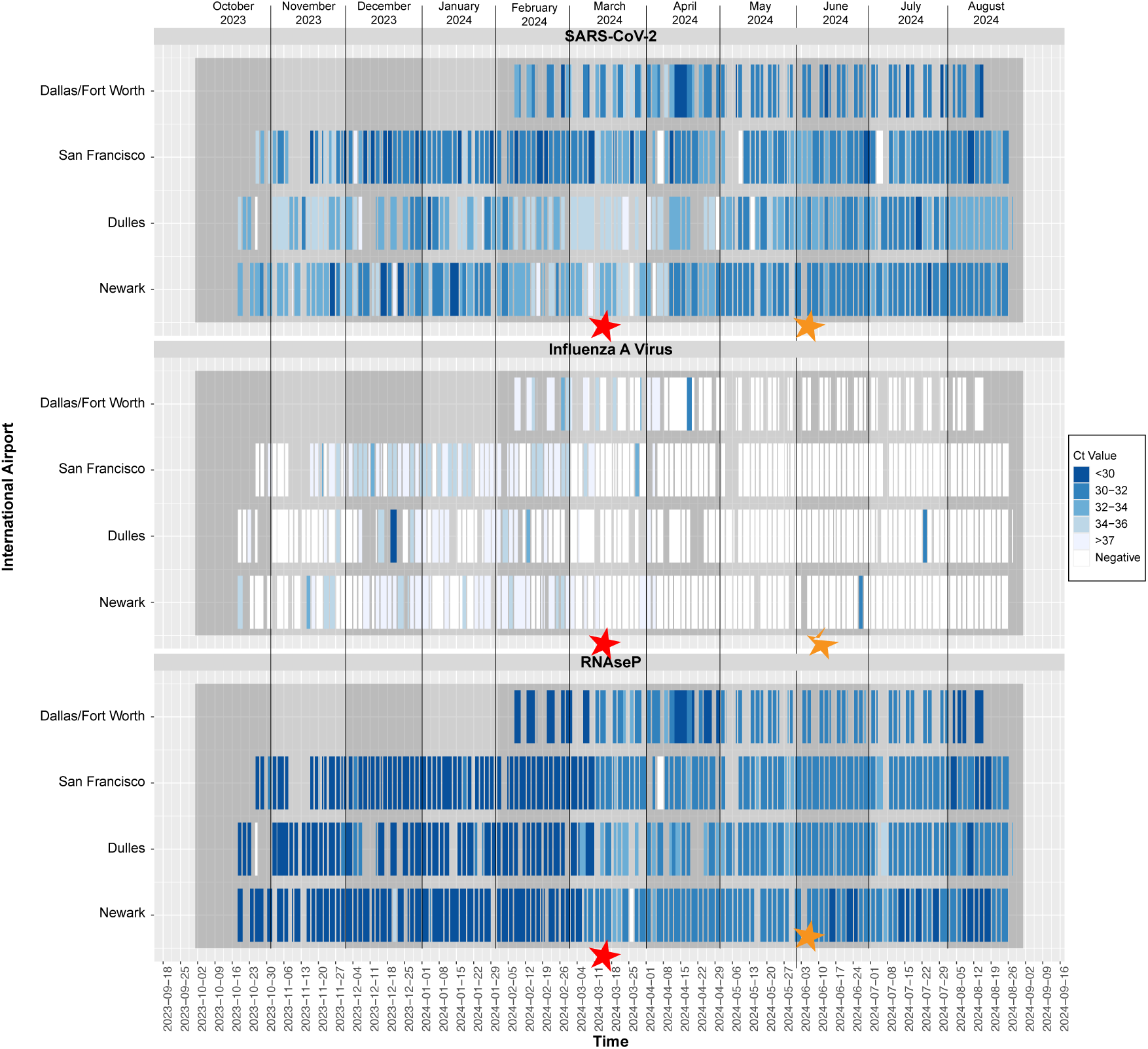
PCR detection from airport air samples. Positive qRT-PCR detections and Ct values of Influenza A, SARS-CoV-2, and RNase P controls captured by ambient air monitoring surveillance from congregate areas in 4 U.S. international airports, October 2023–August 2024. The red star indicates a change of assay; quantitative results before and after this point cannot be directly compared. The orange star indicates when a second sampler was added to the San Francisco airport.

RNA was extracted from 300 µL of eluate using a Maxwell® RSC Viral Total Nucleic Acid Purification Kit (Promega). Molecular detection of SARS-CoV-2, influenza A, and RNase P is described in Supplemental Methods. From October 18, 2023–March 12, 2024, qRT-PCR reactions were performed on a LightCycler® 480 System (Roche) using a TaqMan Fast Virus 1- Step Kit (ThermoFisher) with CDC-modified assays targeting the SARS-CoV-2 nucleocapsid gene (N1, N2) and the influenza A M gene, in duplicate.^19^ Pathogen results were categorized as positive if both Ct values were <40, inconclusive if only one Ct <40, and negative if neither Ct <40. This categorization applied to both SARS-CoV-2 and influenza A assays. On March 12, 2024, the thermocycler was changed to accommodate a multiplex assay (denoted by the red star in Figure 1). For each cartridge, a single multiplex assay measuring the SARS-CoV-2 N1 and influenza A M targets were performed. The human RNase P gene served as an internal control; samples negative for RNase P were excluded from analysis.

For targeted SARS-CoV-2 sequencing, purified total nucleic acid (TNA) was amplified by multiplex RT-PCR (NEB LunaScript Multiplex One-Step RT-PCR) using the ARTIC primer set (IDT ARTIC V5.3.2 NCOV-2019 Panel).^20^ Amplicons were pooled, and libraries prepared with a modified Illumina DNA Prep protocol. Libraries were sequenced on an Illumina NovaSeq 6000, targeting ∼1.5 million paired-end 150 bp reads per sample.

For metagenomic enrichment sequencing, input to library prep was either total nucleic acid (TNA) or clean and concentrated TNA (Zymo Research, Irvine, CA). Extracts were prepared for enrichment using the Illumina RNA for Enrichment Sample Preparation Kit with the Respiratory Virus Oligo Panel, following the manufacturer’s protocol (Illumina, San Diego, CA). Libraries were validated for size and adapter dimer removal on an Agilent TapeStation 4200 D1000 High Sensitivity assay, quantified and normalized using the dsDNA High Sensitivity Assay on a Qubit 3.0 (Life Technologies, Carlsbad, CA), denatured/diluted, and pooled per manufacturer’s instructions. Pooled libraries were sequenced on the Illumina NovaSeq 6000 (v1.5 chemistry) to a target depth of ∼7 million paired-end 150 bp reads per sample. Bioinformatics pipelines and analysis details are provided in Supplemental Data.

For aviation wastewater and traveler nasal swabs, data were generated from samples collected and processed as part of the <u>Traveler Genomic Surveillance (TGS) program</u> from October 18th, 2023-August 27th, 2024. Aviation wastewater was collected from arriving international airplanes at IAD (N=330), and SFO wastewater was collected from triturators (N=161).Triturators are **c**ollection points where waste from multiple flights is discharged and homogenized before being combined with airport terminal wastewater. Travelers volunteered to self-collect anterior nasal swabs on arrival at IAD (N = 16,004) and SFO (N = 30,182). Wastewater samples were concentrated with Nanotrap Microbiome Particles and extracted using a MagMAX Microbiome Ultra Nucleic Acid Isolation Kit (Thermo Fisher). Extracts were tested by digital PCR for SARS-CoV-2 and influenza A/B (Qiagen), and SARS-CoV-2 sequencing was performed as described above.

Upon laboratory receipt, nasal swab samples were heat-inactivated at 70°C for 30 minutes, and total nucleic acids extracted using the MagMAX Viral/Pathogen II Nucleic Acid Isolation Kit (Thermo Fisher). Extracts were tested by qPCR using the TrueMark™ SARS-CoV-2/Flu A/Flu B/RSV Select Panel on a QuantStudio™ 5 qPCR instrument (Thermo Fisher). SARS-CoV-2 sequencing was performed as described above.

This activity was reviewed by CDC, deemed not human subjects research, and conducted in accordance with applicable federal law and CDC policy (45 CFR part 46; 21 CFR part 56).

## Results

During October 18, 2023–August 27, 2024, 463 air monitoring samples were collected and tested by qRT-PCR from four international airports (Figure 1). The RNase P control was positive for 460 of 463 samples (99.1%). Among these 460 RNAseP+ samples, SARS-CoV-2 was detected in 452 (98.3%). Of the remaining eight samples, five were negative and three were inconclusive. When stratified by airport, positive SARS-CoV-2 detections were: SFO: 137 (97.9%), EWR: 124 (98.4%), IAD: 118 (97.5%), and DFW: 73 (100%) (Figure 1).

Among the 460 RNAseP+ samples, 79 were positive for influenza A (17.2%), while 327 were negative and 54 were inconclusive. By airport, influenza A detections were: SFO: 36 (25.7%), EWR: 20 (15.9%), IAD: 10 (8.3%), and DFW: 13 (17.8%). Positive detections of influenza A occurred weekly between January 1 and March 18, 2024, then occurred sporadically through April 22, 2024, after which no further positives were observed (Figure 1).

To compare SARS-CoV-2 and influenza A trends across modalities, data from two airports (SFO and IAD) with overlapping aviation wastewater, airport air, and traveler nasal swab collections were analyzed. Publicly available U.S. clinical respiratory specimen data (https://www.cdc.gov/nrevss/php/dashboard/index.html) were also included for comparison.

Airport air and aviation wastewater had higher weekly positivity rates for SARS-CoV-2 and influenza A than traveler nasal swabs and national clinical samples, reflecting expected differences between population-level (air, wastewater) and individual-level (nasal swab) sampling (Figure 2A). SARS-CoV-2 PCR values from air sampling correlated significantly with aviation wastewater (Figure 2B; Pearson’s p < 0.001), but not with percent positivity in traveler nasal swabs (Figure 2C; p = 0.24), and national surveillance (Figure 2D; p =0.13). Influenza A positivity from air sampling correlated significantly with aviation wastewater (Figure 2E; p = 0.002), traveler nasal swabs (Figure 2F; p < 0.001), and national surveillance (Figure 2G; p < 0.001).

**Figure 2:**
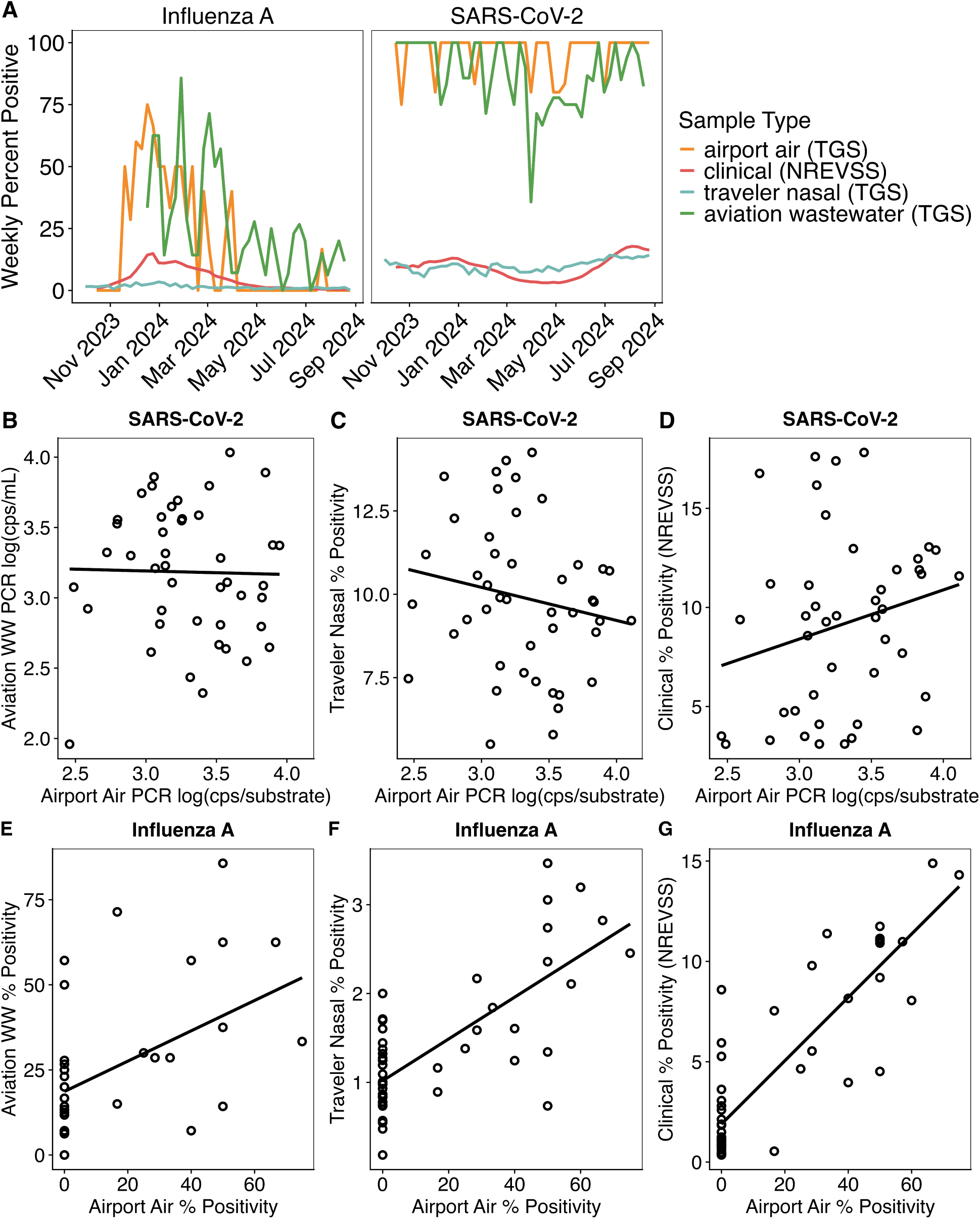
Respiratory virus positivity trends for airport air, aviation wastewater, and traveler nasal swab surveillance. As part of the CDC TGS program, two international airports (SFO and IAD) had overlapping collections for aviation wastewater, airport air, and individual traveler nasal swabs from October 18, 2023-August 27, 2024. Data from these two airports were used to compare virus positivity trends across surveillance modalities. Publicly available data from USA respiratory virus clinical testing (CDC NREVSS) was also included for comparison to national trends. A) Percent positivity for influenza A and SARS-CoV-2 PCR detection compared across surveillance modalities. B-D) Correlation between airport air PCR values for SARS-CoV- 2 and (B) aviation wastewater PCR values, (C) traveler nasal percent PCR positivity, and (D) NREVSS clinical percent positivity. Because percent positivity was near 100% for SARS-CoV-2 in airport air and aviation wastewater samples, PCR values (copies/mL wastewater or copies/substrate for air) were analyzed instead of percent positivity. E-G) Correlation between airport air weekly percent positivity for influenza A and (E) aviation wastewater percent positivity, (F) traveler nasal swab percent positivity, and (G) NREVSS clinical percent positivity.

Of the 460 RNAseP+ air samples, 338 were tested for SARS-CoV-2 sequences; 216 (64%) yielded high-quality genomes (≥70% genome coverage), and 167 (77%) of those contained multiple lineages including BA.2.86, JN.1, JN.1.16, JN.1.67, KP.1, KP.2, KP.3, KP.3.1.1, and XDV. SARS-CoV-2 variant trends in air and aviation wastewater samples were similar at both airports (Figure 3A, 3B). Between January 29 and August 26, 2024, both sample types showed a decline in Omicron JN.1 and an increase in KP.2/KP.3, consistent with contemporaneous national trends.^21^ KP.x variants were detected in aviation wastewater earlier than air samples (42 days earlier for KP.2, 41 days earlier for KP.3).

**Figure 3a, b:**
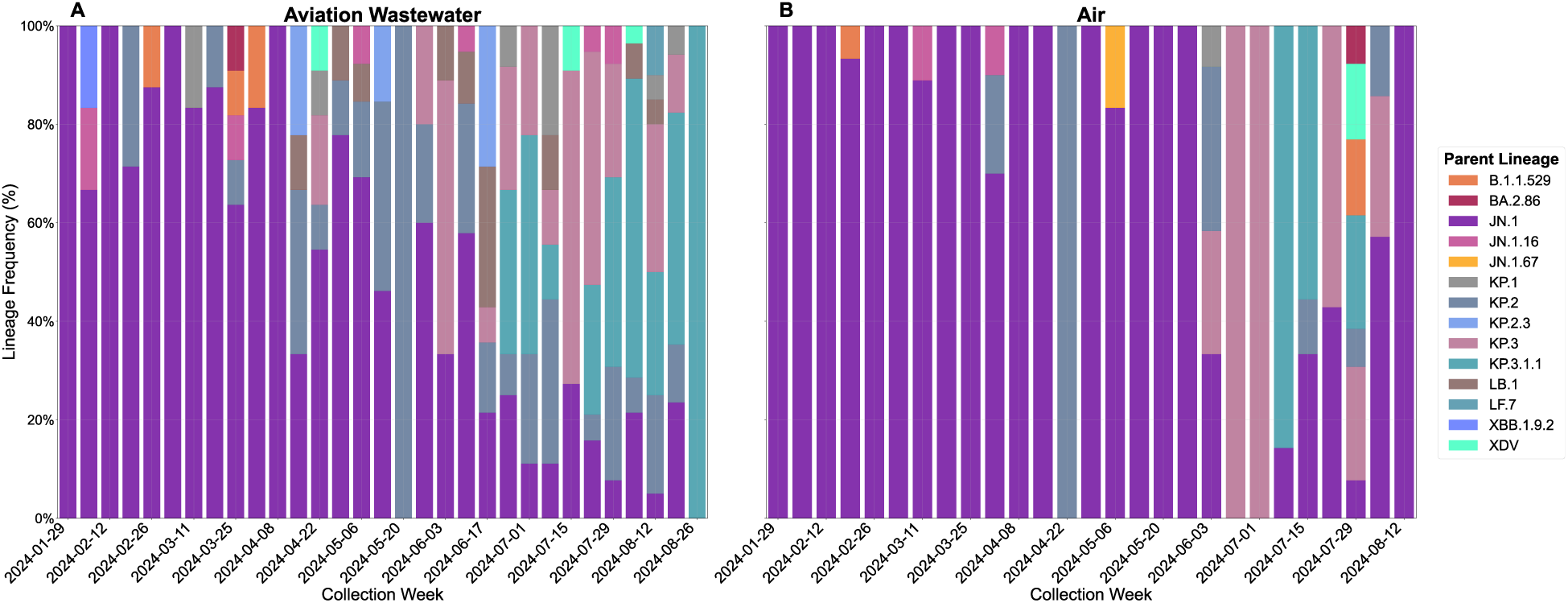
SARS-CoV-2 genomes from wastewater and air samples at two airports (SFO and IAD), samples collected from Jan 29, 2024 to Aug 26, 2024. Genomic sequencing data from **A.** aviation wastewater (SFO-triturator, IAD-airplane) and **B.** Air samples. Aviation wastewater includes both triturator (aggregate airplane lavatory waste) from SFO, and airplane wastewater (individual flight lavatory waste) from IAD.

Metagenomic sequencing with hybrid capture enrichment for respiratory virus genomes was performed on a total of 98 air samples, split into two sequencing runs (Figure 4), using samples that did not overlap with those used for SARS-CoV-2 amplicon sequencing. In the first run (N=52), a mean of 0.04% (95% CI [0.01%, 0.06%]) of total reads per sample mapped to virus targets. After incorporating a TNA concentration step prior to library prep, the second run (N = 46) achieved a mean of 11.8% (95% CI: 7.04 –16.5%) of reads mapping to viral genomes.

**Figure 4:**
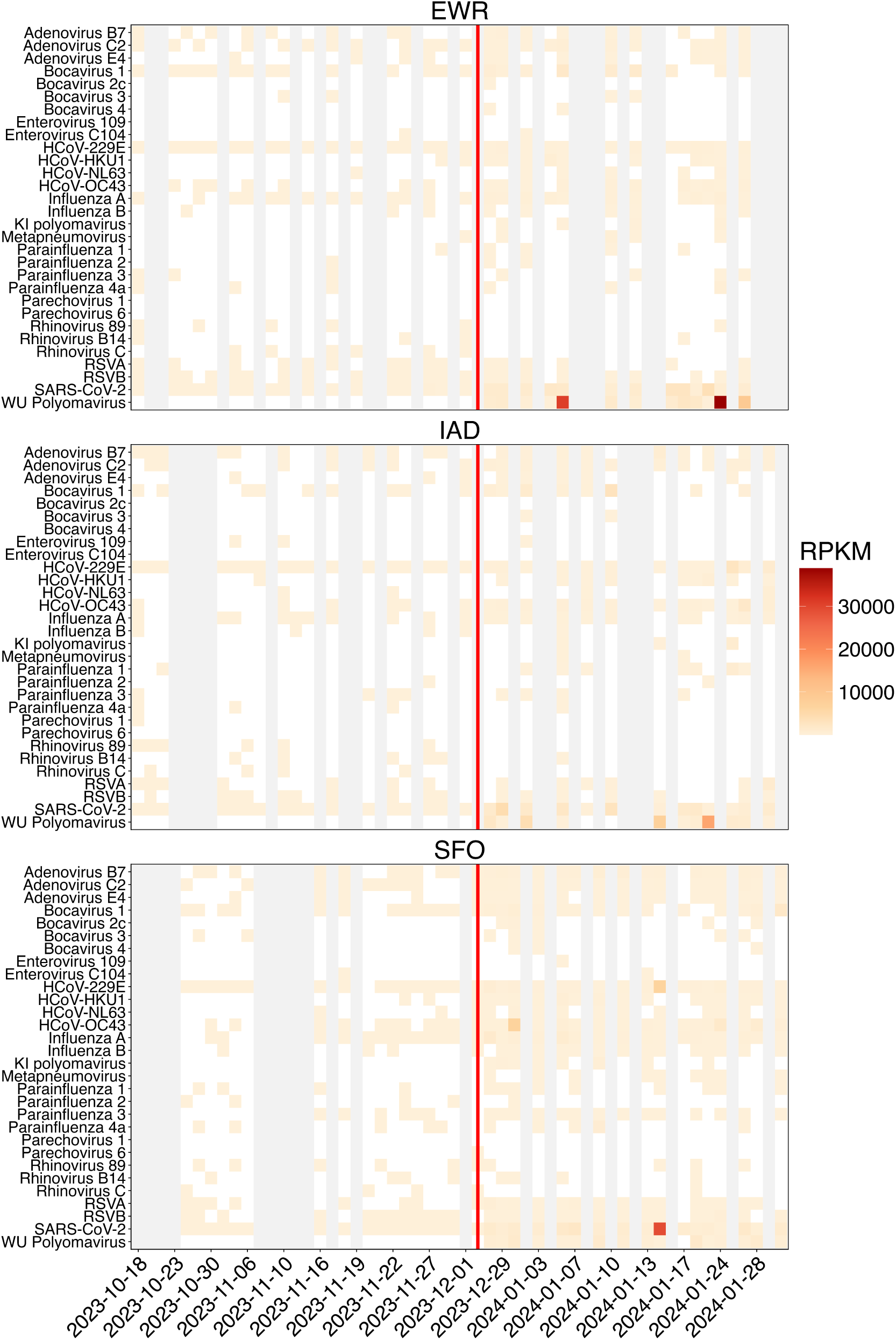
Detection of viral genomes from air samples by metagenomic enrichment sequencing. Nucleic acid extracted from air filters (N=98) collected from three airports (EWR, IAD, SFO) was sequenced using the Illumina Respiratory Viral Oligo hybrid-capture panel for respiratory viruses. Samples to the right of the red line indicate when an RNA concentration step was added to the sequencing library prep protocol. The heatmap shows the normalized abundance of reads mapping to each viral target (RPKM = reads per kilobase per million reads).

Across all samples, reads from 30 viral species were detected. The most frequent were SARS-CoV-2 (100% of samples; mean RPKM 1007, 95% CI: 351-1663), influenza A (57% of samples; mean RPKM 111, 95% CI: 62.7-159), and seasonal coronavirus HCoV-229E (45% of samples; mean RPKM 163, 95% CI: 18.9-306). Influenza B (28%), RSV A (29%), and RSV B (43%) were also detected. For influenza A, 88% (28/32) of PCR-positive samples had sequencing reads mapping to the genome. An additional 27 samples with influenza A reads were PCR- inconclusive (n = 12) or negative (n = 15).

In addition to revealing viral diversity, enrichment sequencing yielded high-quality genomes (≥70% reference coverage at 10× depth) in 4% (2/52) of samples from the first run and 35% (16/46) of samples from the optimized second run. Overall, 25 high-quality genomes were recovered from 98 samples, including SARS-CoV-2 (n = 11), influenza A (n = 4), bocavirus (n = 2), WU polyomavirus (n = 5), and seasonal coronaviruses (n = 3).

## Discussion

The findings of this project support air sampling as a viable option for pathogen surveillance in airports. The data suggests that i) air sampling can serve as a layered or alternative surveillance option, ii) targeted genomic sequencing from air samples provides the resolution needed to identify important variants, and iii) metagenomic enrichment sequencing enables multipathogen detection from air. To our knowledge, this is the first study to demonstrate a correlation between respiratory viruses detected in air samples, aviation wastewater, and nasal swabs from travelers at the same airport, as well as with national clinical respiratory data. Both air and wastewater monitoring offer valuable insights. The strong correlation in observed trends across these two systems underscores their complementary value in a multi-layered surveillance approach, with each capturing signals from distinct yet overlapping populations.

In airport environments, air monitoring offers real-time, surveillance, serving as a crucial interface between emerging threats and community spread. In this project SARS-CoV-2 RNA was detected in nearly all samples (98.3%) compared with 17.2% for influenza A. Influenza A detections were concentrated in the winter months and mirrored national clinical data trends from the 2023-2024 influenza season. The near ubiquitous detection of SARS-CoV-2 underscores the value of genomic sequencing for tracking new variant introductions of highly prevalent pathogens. Sequencing in our study showed JN.1 as the predominant lineage, followed by increases in KP.2 and KP.3 sublineages, consistent with national surveillance data.^21^ In this study, KP.2 and KP.3 lineages were detected substantially earlier in aviation wastewater than in air samples. However early detection was found to be dependent on the sample type when comparing airplane (IAD) and triturator (SFO) wastewater individually against air samples (Supplemental Fig). While some KP.x signals appeared earlier in air than triturator wastewater at specific time points, a controlled comparison across all modalities at the same airport and time would be required to determine whether this reflects true early detection or variation related to low-abundance airborne material.

Genomic sequencing of air sampling has been shown as a highly effective method for pathogen surveillance,^11,12,22^ but methodological considerations remain. Targeted amplicon methods, commonly applied to SARS-CoV-2, have excellent sensitivity, which is valuable for identifying circulating lineages, but interrogate only a narrow set of pathogens. Untargeted metagenomic sequencing captures a broader diversity of pathogens; but the low amount of starting genomic material in air samples presents a technical challenge. Here, we addressed this challenge with hybrid-capture enrichment sequencing, enabling detection of a broad panel of pathogens despite the limitations of input material. Our results show that metagenomic enrichment sequencing increased mapped reads and improved recovery of high-quality genomes from air samples. This approach also detected more influenza A material than PCR, though this may reflect limitations of the specific PCR assay (suggested by the number of inconclusive results) as well as the low sequencing thresholds applied in our study (any read counted as detection).

Our study identified several limitations. First, variations in viral genome reference coverage, due to differing numbers of reads and depth of coverage, may have impacted lineage assignment, particularly in low-biomass air samples. Second, operational constraints meant sampling duration and cartridge change timing were not fully standardized. However, we believe these modest variations did not materially alter the overall findings. Finally, minor temporal uncertainty in signal attribution may have been introduced by the imprecise placement and removal times of the cartridges.

Future research should prioritize enhancing sensitivity by improving air sample collection, nucleic acid recovery, and sequencing efficiency to better detect less abundant pathogens. A parallel focus should be on defining clear detection limits, which will strengthen confidence in genomic data, especially when only limited genomic sequences are recovered.

More broadly, air monitoring technologies currently face technical and operational limitations that require further research and development. Before these systems are widely deployed for biosurveillance, their limits of detection, quantification, and optimal sampling time for pathogens of interest must be rigorously characterized under realistic environmental conditions. In addition, a better understanding of how airflow dynamics, building design, and population density influence sampling efficiency and detection probability would substantially enhance the capability and reliability of these technologies.

Beyond technical considerations, the operational readiness of air monitoring for routine public health use depends on policy, ethical, and implementation factors. These include data interpretability for decision-making, coordination with airport authorities and industry partners, governance around data use, and clear triggers for public health response. As with other environmental surveillance systems, air monitoring is most effective when integrated into established surveillance frameworks rather than deployed as a standalone tool.

Unlike airplane wastewater and traveler nasal swab sampling, which are directly linked to arriving international passengers, air sampling in airport terminals lacks a clear demographic or epidemiological boundary. Air collected in congregate terminal spaces reflects a mixed population that can include airport staff, domestic travelers, and members of the surrounding community, introducing uncertainty regarding attribution to inbound international travel. This limitation reduces the specificity of air sampling for monitoring pathogen importation and must be considered when interpreting variant origin or timing signals relative to other aviation-based surveillance modalities. However, air sampling can provide a complementary layer within a multi-modality framework. Its primary value lies in continuous detection of respiratory pathogens within high-risk indoor environments, capturing signals from individuals who may not contribute to wastewater or participate in voluntary nasal sampling. While air monitoring did not demonstrate earlier lineage detection in all aviation wastewater types in this study, it may offer situational awareness, confirmation of ongoing transmission, and persistence signals during outbreak onset or decline, particularly when other data streams are delayed, unavailable, or operationally constrained.

Air monitoring provides a non-invasive means to track pathogen presence and spread in specific environments. It complements clinical surveillance by capturing signals from large numbers of asymptomatic or mildly symptomatic individuals who would otherwise go undetected. By offering insights into pathogen dynamics, air monitoring can support early threat detection and enable timely public health interventions, even before clinical cases are widely reported. It has been applied in schools^12^ and proposed for long-term care facilities and workplace settings. Air monitoring may also be particularly valuable where wastewater surveillance is impractical, such as in maritime environments, where access to wastewater is limited and where dense populations, close contact, and frequent embarkation/disembarkation increase both transmission risk and the potential for pathogen introduction.^23^

In the future, indoor air detection of viral pathogens in airports or other international ports of entry could become a cornerstone of proactive pandemic preparedness. Continuous or intermittent air sampling, coupled with rapid detection technologies, could provide real-time insights into circulating pathogens. Upon detection of elevated viral loads, targeted mitigation measures, such as increased ventilation^24^, air purification^25^, public health messaging, or resource allocation, could be swiftly launched. This proactive approach, moving beyond reactive clinical case identification, could significantly reduce the risk of disease spread and enhance global health security. Our study highlights air sampling as a feasible, non-invasive strategy for pathogen surveillance. Integrating airport air sampling into existing frameworks can enhance the ability to identify emerging threats and build more resilient, proactive biosecurity systems to address current and future challenges.

## Supporting information

Supplemental Methodology

Air PCR data

Clinical NREVSS data

Enrichment sequencing data

Nasal PCR data

Wastewater PCR data

## Author contributions

DG, CRF, SLO, and DHO contributed to ideas, work, writing, accountability and stewardship. CWP contributed to ideas, work, and stewardship. AR and VJM contributed to work, writing and accountability. BBS contributed to writing, accountability, and stewardship. XQ and PBT contributed to work. TWSA contributed to ideas and work. SB and DJ contributed to work. IR contributed to ideas and accountability.

## Acknowledgements

We are deeply grateful to our partners at XpresCheck for sample collection and support. We thank Amy Schierhorn for program support. We thank Bo Reese and staff at Center for Genome Innovation, Institute for Systems Genomics, University of Connecticut for sequencing support. We thank the CDC Port Health Station staff including Steve Burchell, Erica Sison, Jonathan Georges, Helen MacGregor, Kara Adams, and Ayianna Kennerly. We also thank Miranda Stauss and Eli O’Connor in the O’Connor Labs for cartridge tracking.

## Data Availability

All raw sequencing data is available in the NCBI sequence read archive (SRA) database under CDC’s Traveler-Based Genomic Surveillance Program BioProject PRJNA989177. All other code used to generate figures is available at https://github.com/concentricbyginkgo/TGS-AirMonitoring-Supplemental.

