## Supplemental Methodology for "Environmental air monitoring in international airports: A novel approach for enhanced pathogen surveillance"

**Air sample processing and Quantitative reverse transcription PCR (qRT-PCR)**

ThermoFisher AerosolSense™ cartridge substrates were submerged in 500 μL of 1X phosphate-buffered saline (PBS) and vortexed. RNA was extracted from 300 μL of the PBS eluate using a Maxwell® RSC Viral total nucleic acid purification kit (Promega). Air samples collected from the beginning of the program to March 12, 2024, were tested by qRT-PCR with “Protocol A.” Air samples collected between March 13, 2024 and the end of the program were tested by qRT-PCR with “Protocol B.” Assays used a LightCycler® 480 System (Roche), LightCycler® 96 System (Roche), and a TaqMan Fast Virus 1-Step Kit (Thermo Fisher).

Protocol A: As previously described, the Influenza A virus M gene was tested in duplicate with primers and probes targeting the M gene. SARS-CoV-2 was detected with primers and probes specific for two different targets in the N gene (N1 and N2), and human RNAse P RNA was detected as an internal control.

Protocol B: A multiplex assay for the SARS-CoV-2 N1 target and the Influenza A virus M gene was performed. The N1 target primers and probe were the same as in Protocol A. The IAV M gene primers and probe were designed here (forward primer: 5'-CCMAGGICGAAACGTAYGTICTCTCTATC-3', reverse primer: 5'-TGACAGRATYGGICTTGICTTTAGCCAYTCCA-3', probe: 5'-Cy5-AT YTC GGC T/ZEN/T TGA GGG GGC C/ideoxyI/G /3IABkFQ-3'). The qRT-PCR reaction mix for a 20 μL reaction volume consisted of 3.4 μL of DEPC water, 5.0 μL of TaqMan Fast Virus 1-Step (enzyme), 1.2 μL of 10 μM forward/reverse primers combined with random hexamers, 0.4 μL of 10 μM probes

(N1-FAM and M-Cy5), and 10 µL of the isolated RNA. RNAseP was tested in a separate reaction using the same primers and probe from Protocol A.

Protocol C: Starting July 10, 2024, we changed the IAV target in the multiplex assay for the SARS-CoV-2 N1 target and the Influenza A virus M gene. The N1 target primers and probe were the same as in Protocol A. The new IAV M gene primers and probe were designed here (forward primer: 5'- GGACTGCAGCGTAGACGCTTT -3', reverse primer: 5'-CATCCTGTTGTATATGAGKCCCAT-3', probe: 5'- Cy5-CTMAGYTAT/TAO/TCWRCTGGTGCACTTGCC/3IAbRQSp/). The qRT-PCR reaction mix for a 20 µL reaction volume consisted of 3.4 µL of DEPC water, 5.0 µL of TaqMan Fast Virus 1-Step (enzyme), 1.2 µL of 10 µM forward/reverse primers combined with random hexamers, 0.4 µL of 10 µM probes (N1-FAM and M-Cy5), and 10 µL of the isolated RNA. RNAseP was tested in a separate reaction using the same primers and probe from Protocol A.

**Bioinformatics Pipeline and Analysis**

Raw sequencing data for both SARS-CoV-2 and enrichment sequencing were analyzed using a custom bioinformatics pipeline. Both pipelines were built in Nextflow (v22.04.5). For both pipelines raw sequencing reads were demultiplexed (BCL2fastq; Illumina), reads filtered to those that pass Q30 ≥ 75% (fastqc v0.11.9), adaptors were trimmed (fastp v0.23.2), and human reads were removed (Kraken2 v2.1.2).

For SARS-CoV-2 amplicon sequencing, quality trimmed and filtered reads were aligned to the SARS-CoV-2 reference genome, MN908947.3 (Wuhan-Hu-1), using bowtie2 (v2.4.4). Primers were removed (iVar v1.3.1), alignments were sorted and indexed (samtools v1.15.1), and duplicate reads were marked for high quality alignment (picard v2.27.4 + samtools v1.15.1).

Variant calling and annotation was performed (freebayes v1.3.6, SnpEff v5.0, SnpSift v4.3), and a consensus genome was generated based on quality >100, depth >10, and alternative observations/depth >0.5 (bcftools v1.15.143 INFO/AO / INFO/DP > 0.5 & QUAL > 100 & INFO/DP > 10). SARS-CoV-2 lineages were deconvoluted and frequencies were estimated per sample using Freyja. For consensus sequences, lineages were also identified using Pangolin (v4.3.1). Tool parameter settings were used as default except for fastp (--cut_front --cut_tail --trim_poly_x --cut_mean_quality 30 --qualified_quality_phred 30 --unqualified_percent_limit 40 --length_required 50), freebayes (--ploidy 1 --min-alternate-fraction 0.1, --min-coverage 10), and bcftools (INFO/AO / INFO/DP > 0.5 & QUAL > 100 & INFO/DP > 10).

For metagenomic enrichment sequencing workflow quality trimmed and filtered reads were mapped to a reference database (based on custom selection from NCBI refseq for pathogens on the enrichment panel) using minimap2. Mapping statistics were collected by SAMtools. The pipeline optionally performs IRMA runs for any influenza assemblies, followed by variant calling with freebayes (--ploidy 1, --min-alternate-fraction 0.1, --min-coverage 10) and variant annotation with snpEff. Consensus sequences are generated using bcftools (bcftools view with INFO/AO / INFO/DP > 0.5 & QUAL > 100 & INFO/DP > 10), and Average Nucleotide Identity (ANI) is calculated against references. VCF and consensus sequences undergo filtering based on characterization thresholds (>= 70% consensus reference coverage). Nextclade identifies major lineages for various pathogens including FluA H1/H3/N1/N2, FluB Vic, mpox, SARS-CoV-2, RSVA, and RSVB. Normalized abundance of reads mapping to each viral target is captured as reads per kilobase per million reads (RPKM) compared to viral reference genomes.
